# Effectiveness of Oral Health Education Interventions on Oral Health Literacy Levels in Adults; A Systematic Review

**DOI:** 10.1101/2022.04.04.22273407

**Authors:** Tayebe Ebrahimi, Moshtagh R Farokhi

**Author notes:** Correspond to: Tayebe Ebrahimi.

## Abstract

**Background:** Oral health literacy within the construct of health literacy may be instrumental in decreasing oral health disparities and promoting oral health. Even though current research links oral health literacy to oral health knowledge and education, the impact of educational intervention on oral health literacy remains controversial. We aimed to identify effective health education interventions delivered with a focus on oral health literacy.

**Methods:** An electronic systematic search in PubMed, Scopus, Web of Science and Cochrane library and gray literatue was performed for relevant studies (1995-2021). Experimental study designs of randomized controlled trials, non-randomized controlled trials, and quasi-experimental studies in which adults aged 18 years or older, male, or female (participants) trained under a health education intervention (intervention) were compared with those with no health education or within the usual care parameters (comparison). An assessment of oral health literacy levels (outcome) were included according to the PICO question. The search was conducted by applying filters for the title, abstract and methodological quality of the data, and English language. Study screening, extraction and critical appraisal was performed by two independent reviewers. Data was extracted from the included studies whereas a meta-analysis was not possible since findings were mostly presented as a narrative format.

**Results:** Eight studies out of the 2783 potentially eligible articles met the selection criteria for this systematic review. The aim of interventions in these studies was 1) improving oral health literacy as the first outcome or 2) improving oral health behavior and oral health skills as the first outcome and assessing oral health literacy as the second outcome. The strength of evidence from the reviewed articles was high and there was an enormous heterogeneity in the study design, OHL measurement instruments and outcomes measure. Interventions were considerably effective in improving oral health literacy.

**Conclusion:** Health education that is tailored to the needs and addresses patients’ barrier to care can improve their oral health literacy level.

## Background

According to the scientific evidence on health inequalities, lower level of education is associated with lower health outcomes [1]; however, the mechanism of this association is not yet well understood [2]. Recently, health literacy has been at the intersection between educational achievements and health outcomes. Health literacy is the capacity to obtain, understand and process health information that enables an individual to make decisions regarding services needed by utilizing their cognitive and social skills [3]. Individuals are more likely change their behavior when they achieve suitable amount of knowledge, self-efficacy, and the sense of controlling their health consequences [4]. Health literacy (HL) is increasingly realized as an essential contributor to overall health outcomes where a low level of health literacy is identified as a major contributor to health inequality [5]. Health literacy plays an important role in enabling access to health care that ultimately improves individual and community health outcomes.

Oral health literacy (OHL) as a subset of health literacy has also proven to decrease oral health disparities in promoting promote oral health. Even though oral health professionals intentionally focus on educational approaches, there are occasions that patients and their families my misunderstand the message. Misunderstanding of the oral health instructions and inability to listen, read and critically analyze verbal and written information as brochures is a consequence of low OHL [6]. Oral health literacy is a global phenomenon as related to oral health disparities.

Socioeconomic factors, oral health attitudes, beliefs and practices can either improve or diminish oral health outcomes. A more advanced individual oral health status yields to a higher score of OHL or improved adopted oral health-related behaviors [7]. In contrast, individuals with lower OHL, experience higher levels of oral disease including dental caries and periodontal disease that leads to tooth extractions and loss of function.

Numerous tools are currently available to assess OHL with their origins derived from health literacy assessing tools. For example, the tool of Rapid Estimate of Adult Literacy in Dentistry [8] has been adopted from the Rapid Estimate of Adult Literacy in Medicine [9]. In a similar fashion, the Test of Functional Health Literacy in Dentistry [10] is an adaptation of the Test of Functional Health Literacy in Adults [11]. However, other Oral health literacy tools have been designed specifically for dentistry such as OHL Adults Questionnaire (OHL-AQ), [12]. Health education intervention improves health through the gaining of knowledge and learning skills, that leads to actions conductive to health. However, pure acquisition of health knowledge does not lead to correct health behaviors. Average intervention comprises of attempts to provide oral health information accessibility to individuals and populations with limited literacy. Examples include the use of PowerPoints, pictographs, charts, posters, and electronic media to deliver health information that is easier to read and understand.

Although current studies show that oral health literacy is associated with oral health knowledge and education, the effect of educational intervention on oral health literacy is controversial [13]. Other studies suggested oral health hands-on training programs for people with lower education and literacy skills [14]. This systematic review identifies the best evidence on effectiveness of health education interventions on the level of oral health literacy in adults.

## Methods

### Data Sources

We searched PubMed, EMBASE, Scopus and Web of Science. We formulated a combination of MeSH and non-Mesh terms and keywords in consultation with two professional Health Sciences Librarians to make sure of completeness (Appendix 1). These searches included all identified multiple terms for health education intervention (e.g., health program, health intervention) and oral health literacy (e.g., health literacy in dentistry).

Google scholar was also searched with these keywords manually to retrieve the gray literature. A search on gray literature was also conducted. The references of the included studies were considered for more relevant studies. The publication dates were restricted from 1995-2021. The initial search was conducted in September 2021 and updated in February 2022.

### Study Selection

This review included studies that involved the population of adult age group of 18 years and older without any gender restrictions. Any educational intervention reported to improve oral health was included with the aim to either change a predefined aspect of oral health status/outcomes or address oral health literacy. Both population and individual level interventions were included. Non-intervention or usual care was considered as comparator/control group.

The studies in English language that fulfilled the following inclusion criteria according to the PICO acronym. We included randomized clinical trials, non-randomized clinical trials, pre and post quasi-experimental and controlled before and after studies. Studies that demonstrated outcome measures related to oral health literacy were included. Case reports, case series, cross sectional studies, reviews, letters to the editor, studies with animals, technical notes, and studies were not in English language were excluded.

The proposed systematic review was conducted in accordance with the methodology for systematic reviews of qualitative evidence. The review protocol was registered with PROSPERO (registration number CRD42021275828). All studies meeting the selection criteria were assessed for their methodological quality.

### Data synthesis

The screening was done independently by two reviewers. Cohen’s Kappa was used to measure the agreement between researchers that was considered in agreement (Kappa = 0.86). Studies were eligible to include in this review if they met the following parameters: clinical trials and before after quasi-experimental studies that evaluated the effects of education intervention on the level of oral heath literacy. Moreover, evaluation of OHL had to be performed through validated instrument.

First, the duplicated retrieved search results were identified and excluded. We employed endnote software for identifying duplicate studies. We screened the titles and the abstracts of the papers to exclude the irrelevant studies. Search results were categorized into three categories of included, excluded and unclear. Later, the full texts of the included and unclear studies were reviewed for final inclusion. Any disagreement was discussed and resolved between the two reviewers. Any further disagreements between the two reviewers were to be resolved through a third reviewer.

We extracted data from eligible articles based on study characteristics (study title, authors, date of publication, study design, inclusion/exclusion criteria of patients, sample size); baseline data (education intervention model, oral health literacy assessment tool) and clinical outcomes (changes in the level of OHL). We systematically reviewed studies comprising adults trained under a health education intervention program and underwent assessing their oral health literacy level. Within this scoping review, we adhered to the Preferred Reporting Items for Systematic Reviews and Meta-Analyses (PRISMA) statement 2020 recommendations provided by Liberati [15], (Figure 1).

**Figure 1:**
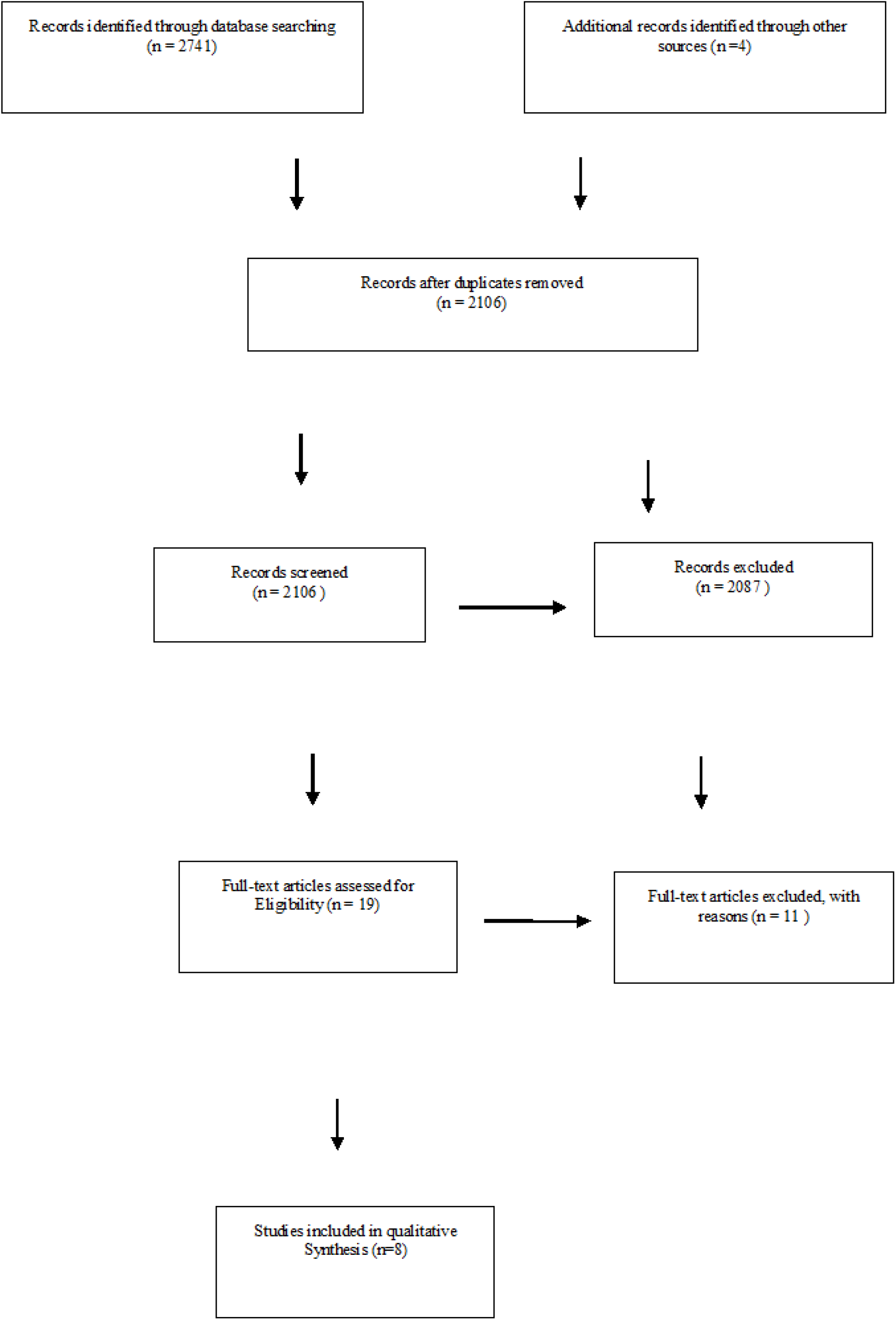
PRISMA diagram

We forbore performing a meta-analysis due to finding that studies lacked the same interventions, or standardized approaches to outcomes measurements. Therefore, we performed a qualitative analysis. In total, eight intervention studies were recognized. This review consisted of three randomized controlled trials and five quasi experimental pretests to post-tests interventional studies; all study designs were rated good standard regarding quality appraisal.

## Results

This systematic review retrieved 2783 journal article. A total of 1329 articles remained after the removal of duplications. 2087 studies excluded during title and abstract screening as they did not meet inclusion criteria. 19 studies were included for full text review and assessment. Eight studies were included in the final synthesis. Table 1 demonstrates reasons for study exclusion. Our sample size was between 50 and 478 subjects where1610 subjects were included.

**Table 1:** Reasons for exclusion of Articles

| Author | Study Theme | Study Design | Reasons for Exclusion |
| --- | --- | --- | --- |
| Parker (2012) | Oral health education workshops | Protocol of a Randomized control trial | Results of intervention is not published |
| Villela (2017) | Oral health literacy education consisted of information about nutrition and oral hygiene for children <2 years old | Randomized control trial | Knowledge score was assessed instead of OHL scores |
| Rani (2019) | Health education for school students about health teeth and gingiva and symptoms of dental and gingival disease | Quasi experimental pre-post test | Tooth and gingival drawing scores were assessed instead of OHL scores |
| Ueno (2013) | Health education for dental students about health teeth and gingiva and symptoms of gingival disease | Quasi experimental pre-post test | Drawing scores was assessed instead of OHL scores |
| Naito (2007) | Searching for health information through internet and benefits of tooth brushing for prevention of gingival diseases were presented | Quasi experimental pre-post test | OHL was not assessed |
| Stein (2018) | Recommended communication techniques were presented | Randomized Controlled Trial | OHL was not assessed |
| Jeelani (2017) | Oral health literacy presentation on HIV | Quasi experimental pre-post test | Difference in knowledge before and after the intervention was assessed |
| Lisboa (2020) | Information about how to respond to traumatic dental injuries was presented | Quasi experimental pre-post test | Knowledge of traumatic dental injuries was assessed |
| Vilella (2017) | A health education intervention contained information about the eating habits and oral hygiene of children younger than 2 years | Randomized controlled trial | Knowledge scores was assessed |
| Khudanov (2018) | Dental plaque removal was trained using the Qscan device | Randomized controlled trial | oral health knowledge, behavior, and attitude were evaluated |

The included reports took place in five countries: Australia [16], Canada [17], Iran [18], The United States of America [19-21], Malaysia [22], and China [23]. The earliest report was from 2014 [21], and the most recent article was from 2021 [23] (Table 2).

**Table 2:**
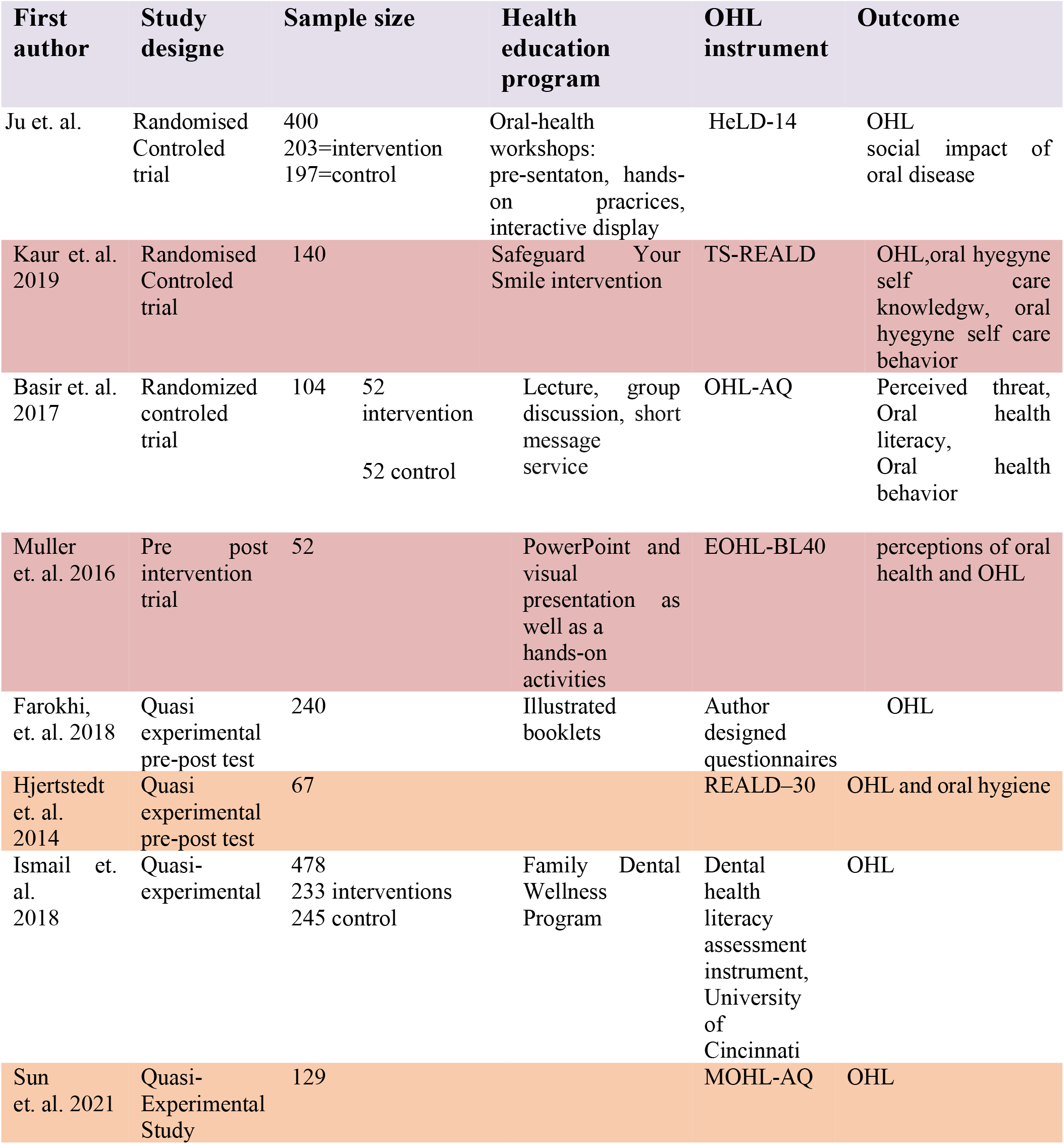
Reasons for Inclusion

One study was blinded randomized control trial [18], two studies were non-blinded, randomized control trial [16-17], one study was non-randomized quasi experimental with a control group [22], and four were quasi experimental reports without a control group [19-21, 23].

### Intervention strategies and oral health literacy tools

Health education intervention and outcome and measurement of oral health literacy varied considerably in the included reports for this systematic review where all the studies provided educational intervention to adult participants. Assessing oral health literacy before and after intervention was a primary outcome of six studies [16, 19-22, 23] and the secondary outcome in two studies [17-18].

One health education intervention included five interactive workshops presented by Indigenous staff. It encompassed information about dental diseases, getting familiar with dental clinic, access to dental services and patient rights. Each workshop session lasted one and a half hours with a follow up in one year. They utilized the HeLD 14 for assessing oral health literacy [16].

A randomized controlled trial “Safeguard Your Smile intervention (SYS)” program conducted by the lead researcher consisted of 1) understanding the risk factors of dental plaque and gingivitis and effects of oral-hygiene-self-behavior on health, 2) demonstrating tooth brushing and flossing tools and skills, 3) encouraging the planning for oral hygiene daily routine, 4) tracking the progress of oral hygiene self-care every day for three month, and 5) follow up calls to reinforce participant behavior. The TS-REALD was used for assessing oral health literacy [16].

Other educational interventions consisted of one in-person lecture and group discussion session. During this session, caregivers received appropriate tips about on how to breastfeed at night, and how to brush or clean children’s teeth by demonstrating the correct methods of tooth brushing. To keep the participants motivated, brief intervention, and educational short message service (SMS) reminders were sent every two weeks for 6 months. They utilized the OHL tool from relevant literature [18].

One health education intervention consisted of a 50-minute visual demonstration as well as PowerPoint presentation and hands-on activities conducted by a principal investigator. Participants learned tooth brushing and flossing techniques. The OHL questionnaire in this study was EOHL-BL40 [19].

Another intervention was to empower and equip participants with knowledge and skills to advance access to oral health care, promote preventive care, oral health self-care, dietary counseling, and emphasis on the integration of oral health to systemic health by the dental student and faculty to medical and nursing students, refugee patients and community members. At these sessions, dental and dental hygiene students utilized an illustrated booklet to further oral health literacy capacity by providing information in a 15-20 min presentation. A pre and post author created questionnaire centered at caries, periodontal and oral cancer risk assessment was administrated to participants [20].

One health education program consisted of a five 2-hour visits. OHL, oral hygiene, impact of fluoride and saliva and medication, dietary approaches to oral health and oral-systemic connections were topics of these visits. First year Dental students scheduled the visits over a six-month period. REALD–30 was used as OHL questionnaire [21].

Another health education intervention approach addressing a cognitive component, psychomotor component, and an attitude component. Participants were required to make visits every six months for the duration of three years where they took part in a one-hour oral health literacy session conducted by dentists or dental therapists for a three-year follow-up period. The OHL questionnaire in this study was based on Ludke R’s Dental health literacy assessment instrument [22].

One program was a 30 min course with “easy to read” written materials that was delivered by and PowerPoint slides (contained guidelines published by Daghio et al.). They used Mandarin version of the oral health literacy adult questionnaire (MOHL-AQ) or [23].

### Methodological Quality Assessment

We applied the latest version of risk of bias 2 (RoB 2.0) checklists for quality appraisal of randomized clinical trial studies as well as the JBI critical appraisal checklist for quasi-experimental studies. The criteria for quality in Rob2 tool include randomize process, deviation from intended intervention, missing outcome data, measurement of the outcome and selection of the reported result. The criteria for quality in JBR critical appraisal tool include clarity of the cause, similarity of participants, receiving similar care, existence of control group, existence of multiple measurements before and after intervention, completion of follow up, similarity of outcome measurement, reliability of outcome measurement, appropriate analysis. We assessed three studies using Rob2 and five using JBI critical appraisal checklist. The results of quality assessment of included studies are displayed as Appendix 2 and 3. Achieving overall + grade in Rob2 and p more than 6 point in JBI was considered as high quality study. All included studied had high methodological quality.

The randomized control trial studies explained sufficient randomization, intervention, outcome measurement, and proper statistical analysis. Ju et al. [16], analyzed collected data under three different scenarios. For the first scenario, all samples were analyzed regarding the original intervention. For the second scenario, any intervention group participant who didn’t attend a workshop session was moved to control group. In the third scenario, participants who didn’t attend any workshop session were excluded. According to crude data analysis, there was no significant difference for oral health literacy between the two groups undertaking the three scenarios. Multiple imputation (MI) was used to replace missing data and after MI oral health literacy improved under the second scenario.

The study of Kaur et al. [17], reported significant increase in the OHL scores after SYS intervention. Basir et al. [18], found a significant difference between two groups in term of oral health literacy. In the study of Muller et al. [19], OHL scores were significantly higher after oral health education program where the OHL scores were significantly higher in participant’s native language compared to English language.

Results showed a significant increase in percentage of knowledge gained post OHL questionnaire intervention as compared to the baseline information in the study by Farokhi et al. [20]. There was a significant higher OHL scores following completion of health education visits than the baseline scores in the study of Hjertstedt et al. [21].

Ismail et al. [22], showed that family dental wellness program significantly increased oral health literacy scores in intervention group in compare with control group. Sun et al. displayed that OHL scores significantly improved using easy-to-read material [23].

## Discussion

Oral health literacy is a concept that includes social determinants of health such as education and socioeconomic status. Previous studies have reported that higher level of education leads to higher oral health literacy [14, 25]. Statistically significant relationships have been observed between oral health literacy level and oral health status [26]. This systematic review aimed to explore the effect of health education interventions on the level of oral heal literacy from 1995 until February 2022.

### Systemic Reviews of Health Literacy

Although we found a limited number of systematic reviews which considered oral health literacy as an outcome, the systematic reviews of health literacy were numerous. A Systematic review that assessed effectiveness of HL interventions in European union included 23 studies. Most of these studies focus on functional HL, fewer considered interactive or critical HL and authors couldn’t draw a firm conclusion due to the low quality of the included studies [27].

Another systematic review search assessing efficacy of interventions to improve HL and health behaviors yielded 22 studies. Results showed that HL intervention could improve aspects of HL and behavioral outcomes in most studies [28]. One systematic review explained that complex interventions targeting people with limited literacy can improve health knowledge and self-efficacy [29]. Another systematic review explained mixed results for health intervention in people with low literacy but couldn’t draw a conclusion regarding the study design [30].

Health education interventions delivered with a focus on oral health literacy

### Target Population

Participants populations varied in our eight included studies. Participants were indigenous Australian adults [16], Punjabi immigrants [17], women who were referred to a health care center [18], refugee population [19-20], older adults who resided in independent or assisted living apartments [21], and eligible children with caries referred to two government dental clinics [22]. In one study participants were from different populations of refugees, community members, and medical and nursing students [20] and another utilized bibliographic records [23].

### Types of Interventions

A wide range of educational interventions were investigated. One approach was an oral health literacy intervention [16], while another utilized the “Behavior Change Wheel” which was applied to improve the oral hygiene self-care behavior [17]. Another intervention observed the children’s oral health as well as standard well baby care which contained knowledge about immunization and child growth where the participants received Short Messages Service reminders of health education every two weeks [18]. An intervention focused on educational approach to proper toothbrushing and flossing techniques [19]. One intervention was an illustrative booklet used to inform participants with variable levels of oral health literacy [20]. Oral health and oral hygiene education was another intervention [20-21]. Participants in another intervention were empowered to improve and maintain oral health of their children under the Family Dental Health Program [22], (Table 2).

### Intervention Significance

Overall, there were no systematic review studies that assessed OHL after health education intervention, however, some studies investigated HL. In a randomized control trial by Zhuang et al. [31], the results indicated that receiving health education SMSs every week for one year, significantly increased health literacy scores of the adults as compared with control group. This finding was also confirmed as the studies done by Basir et al. and Ismail et al. In another randomized trial by Otsuka-Ono et al. [32], educational sessions were designed to provide pregnant women with child immunization information developed through group and individual interview and the use of a guidebook as a communication tool. Health literacy scores were significantly higher in intervention group which again confirmed the results of studies by Basir [18], and Ismail [22].

In a HL intervention, HIV positive people received education program consisted of using computer and internet for safe eHealth searching. HL scores of immediately post intervention was significantly higher but after 3-months follow-up, there were no significant difference in compare with pretest scores [33]. An education intervention designed to improve decision making and HL among older Australian, there were no significant difference between the group which received complementary medicine using website or DVD and a booklet versus a booklet only [34]. McCaffery et al. evaluated the effectiveness of a HL program in adults with low literacy in a randomized clinical trial study. Participants received a standardized program which contained language, literacy and numeracy or HL program. They received the aforementioned items in standard program as well as embedded health content whereas HL changes were not significantly different when the two groups were compared [35]. Jervelund et al. [24], investigated healthcare-seeking behavior before and after an information intervention among immigrants, results showed that actual healthcare use of immigrants didn’t change after receiving either a course or written information on the Danish healthcare system of intervention.

### Aims of interventions

The aim of interventions in these studies were to 1) improve oral health literacy as the first outcome, 2) improve oral health behavior and 3) improve oral health skills as the first outcome and assessing oral health literacy as the second outcome. Interventions were mainly presented in the form of individual or group workshops. Although validated tools were applied to measure OHL, six different tools were used in the reviewed studies. Four studies utilized previously designed oral health questionnaire [17, 19, 21-22], and the other four studies designed specific questionnaire for their studies [16, 18, 20].

### Conclusion

Among the three randomized controlled trials included in this study, two were targeting mothers of pre-school children and both reported significant difference between baseline and post intervention oral health literacy scores. OHL level can influence processing and understanding preventive oral health information; therefore, the OHL of caregivers impairs oral health status of their children. Reviewing of these eight studies suggested that health education programs could foster oral health literacy scores in adults.

This systematic review finds that validated OHL instruments are instrumental in measuring oral health literacy for future studies. Additionally, further studies should evaluate oral health literacy as their first outcome.

### Limitations

A limitation of our study was that limited full text articles were included from the five academic databases.

## Data Availability

All relevant data are within the manuscript and its Supporting Information files.

## Appendix 1: Search Strategy

| Data base | Search strategy |
| --- | --- |
| PubMed | ("health education" OR (Education AND Health) OR "Community Health Education" OR Workshops OR Workshop OR "Training program" OR "Training programs" OR "training programme" OR (Training AND program) OR (Training AND programs) OR (training AND programme) OR "Educational activities" OR "Educational activity" OR (Educational AND activities) OR (Educational AND activity) OR "Educational workshop" OR (Educational AND workshop) OR "Educational workshops" OR (Educational AND workshops) OR "Educational program" OR "Educational programs" OR "Educational training" OR "Educational trainings" OR (Educational AND training) OR "Literacy program" OR "Literacy programs" OR (Literacy AND program) OR (Literacy AND programs) OR "health literacy workshop" OR "health literacy workshops" OR ("health literacy" AND workshop) OR ("health literacy" AND workshops) OR "health educational intervention" OR "health educational interventions" OR "health educational workshop" OR "health educational workshops" OR "health educational program" OR "health educational programs" OR "health educational activity" OR "health educational activities" OR "health educational training" OR "health educational trainings" OR "health education intervention" OR "health education interventions" OR "health education workshop" OR "health education workshops" OR "health education program" OR "health education programs" OR "health education activity" OR "health education activities" OR "health education training" OR "health education trainings" OR "health literacy intervention" OR "health literacy interventions" OR ("health literacy" AND intervention) OR ("health literacy" AND interventions) OR "health literacy program" OR "health literacy programs" OR ("health literacy" AND program) OR ("health literacy" AND programs) OR "health literacy activity" OR "health literacy activities" OR ("health literacy" AND activity) OR ("health literacy" AND activities) OR "health literacy training" OR "health literacy trainings" OR ("health literacy" AND training) OR ("health literacy" AND trainings) OR (Education AND "oral health") OR "oral health education" OR (workshop AND "oral health") OR "oral health workshop" OR (workshops AND "oral health") OR "oral health workshops" OR (workshop AND dental) OR (workshop AND "dental health") OR "Dental Health workshop" OR (workshops AND "dental health") OR "Dental Health workshops" OR (Education AND "dental health") OR "Dental Health Education" OR "oral health program" OR "dental health program" |
|  | OR (program AND “oral health”) OR (program AND “dental health”) OR “oral health training” OR “dental health training” OR (training AND “oral health”) OR (training AND “dental health”) OR “oral health activity” OR “dental health activity” OR “oral health activities” OR “dental health activities” OR (activity AND “oral health”) OR (activity AND “dental health”) OR (activities AND “oral health”) OR (activities AND “dental health”) OR “oral health intervention” OR “oral health interventions” OR “dental health intervention” OR “dental health interventions” OR “health program” OR “health training” OR “health activity” OR “health programs” OR “health workshop” OR “health workshops” OR “health trainings” OR “health activities” OR “health intervention” OR “health interventions” OR “Patient Education as Topic” OR (Education AND Patient) OR “Patient Education” OR “Education of Patients”) AND (“oral health literacy” OR “dental health literacy” OR “literacy in dentistry” OR “dental health literate” OR “oral health literate” OR (“health literacy” AND “oral health”) OR (“health-related literacy” AND “oral health”) OR (“health Literacy” AND “dental health”))<br>1228 |
| Web of science | (ALL=“health education” OR ALL=(Education AND Health) OR ALL=“Community Health Education” OR ALL=Workshops OR ALL=Workshop OR ALL=“Training program” OR ALL=“Training programs” OR ALL=“training programme” OR ALL=(Training AND program) OR ALL=(Training AND programs) OR ALL(training AND programme) OR ALL=“Educational activities” OR ALL=“Educational activity” OR ALL=(Educational AND activities) OR ALL=(Educational AND activity) OR ALL=“Educational workshop” OR ALL=(Educational AND workshop) OR ALL=“Educational workshops” OR ALL=(Educational AND workshops) OR ALL=“Educational program” OR ALL=“Educational programs” OR ALL=“Educational training” OR ALL=“Educational trainings” OR ALL=(Educational AND training) OR ALL=“Literacy program” OR ALL=“Literacy programs” OR ALL=(Literacy AND program) OR ALL=(Literacy AND programs) OR ALL=“health literacy workshop” OR ALL=“health literacy workshops” OR ALL=(“health literacy” AND workshop) OR ALL=(“health literacy” AND workshops) OR ALL=“health educational intervention” OR ALL=“health educational interventions” OR ALL=“health educational workshop” OR ALL=“health educational workshops” OR ALL=“health educational program” OR ALL=“health educational programs” OR ALL=“health educational activity” OR ALL=“health educational activities” OR ALL=“health educational training” OR ALL=“health educational trainings” OR ALL=“health education intervention” OR ALL=“health education interventions” OR ALL=“health education workshop” OR ALL=“health education workshops” OR ALL=“health education program” OR ALL=“health education programs”) OR ALL=“health education activity” OR ALL=“health education activities” OR ALL=“health education training” OR ALL=“health education trainings” OR ALL=“health literacy intervention” OR ALL=“health literacy interventions” OR ALL=(“health literacy” AND |
|  | intervention) OR ALL=(“health literacy” AND interventions) OR ALL=“health literacy program” OR ALL=“health literacy programs” OR ALL=(“health literacy” AND program) OR ALL=(“health literacy” AND programs) OR ALL=“health literacy activity” OR ALL=“health literacy activities” OR ALL=(“health literacy” AND activity) OR ALL=(“health literacy” AND activities) OR ALL=“health literacy training” OR ALL=“health literacy trainings” OR ALL=(“health literacy” AND training) OR ALL=(“health literacy” AND trainings) OR ALL=(Education AND “oral health”) OR ALL=“oral health education” OR ALL=(workshop AND “oral health”) OR ALL=“oral health workshop” OR ALL=(workshops AND “oral health”) OR ALL=“oral health workshops” OR ALL=(workshop AND dental) OR ALL=(workshop AND “dental health”) OR ALL=“Dental Health workshop” OR ALL=(workshops AND “dental health”) OR ALL=“Dental Health workshops” OR ALL=(Education AND “dental health”) OR ALL=“Dental Health Education” OR ALL=“oral health program” OR ALL=“dental health program” OR ALL=(program AND “oral health”) OR ALL=(program AND “dental health”) OR ALL=“oral health training” OR ALL=“dental health training” OR ALL=(training AND “oral health”) OR ALL=(training AND “dental health”) OR ALL=“oral health activity” OR ALL=“dental health activity” OR ALL=“oral health activities” OR ALL=“dental health activities” OR ALL=(activity AND “oral health”) OR ALL=(activity AND “dental health”) OR ALL=(activities AND “oral health”) OR ALL=(activities AND “dental health”) OR ALL=“oral health intervention” OR ALL=“oral health interventions” OR ALL=“dental health intervention” OR ALL=“dental health interventions” OR ALL=“health program” OR ALL=“health training” OR ALL=“health activity” OR ALL=“health programs” OR ALL=“health workshop” OR ALL=“health workshops” OR ALL=“health trainings” OR ALL=“health activities” OR ALL=“health intervention” OR ALL=“health interventions” OR ALL=“Patient Education as Topic” OR ALL=(Education AND Patient) OR ALL=“Patient Education” OR ALL=“Education of Patients”) AND (ALL=“oral health literacy” OR ALL=“dental health literacy” OR ALL=“literacy in dentistry” OR ALL=“dental health literate” OR ALL=“oral health literate” OR ALL=(“health literacy” AND “oral health”) OR ALL=(“health-related literacy” AND “oral health”) OR ALL=(“health Literacy” AND “dental health”)) |
| Scopus | (ALL(“health education”) OR ALL((Education AND Health)) OR ALL(“Community Health Education”) OR ALL(Workshops) OR ALL(Workshop) OR ALL(“Training program”) OR ALL(“Training programs”) OR ALL(“training programme”) OR ALL((Training AND program)) OR ALL((Training AND programs)) OR ALL((training AND programme)) OR ALL(“Educational activities”) OR ALL(“Educational activity”) OR ALL((Educational AND activities)) OR ALL((Educational AND activity)) OR ALL(“Educational workshop”) OR ALL((Educational AND workshop)) OR ALL(“Educational workshops”) OR |
|  | ALL((Educational AND workshops)) OR ALL("Educational program") OR ALL("Educational programs") OR ALL("Educational training") OR ALL("Educational trainings") OR ALL((Educational AND training)) OR ALL("Literacy program") OR ALL("Literacy programs") OR ALL((Literacy AND program)) OR ALL((Literacy AND programs)) OR ALL("health literacy workshop") OR ALL("health literacy workshops") OR ALL(("health literacy" AND workshop)) OR ALL(("health literacy" AND workshops)) OR ALL("health educational intervention") OR ALL("health educational interventions") OR ALL("health educational workshop") OR ALL("health educational workshops") OR ALL("health educational program") OR ALL("health educational programs") OR ALL("health educational activity") OR ALL("health educational activities") OR ALL("health educational training") OR ALL("health educational trainings") OR ALL("health education intervention") OR ALL("health education interventions") OR ALL("health education workshop") OR ALL("health education workshops") OR ALL("health education program") OR ALL("health education programs") OR ALL("health education activity") OR ALL("health education activities") OR ALL("health education training") OR ALL("health education trainings") OR ALL("health literacy intervention") OR ALL("health literacy interventions") OR ALL(("health literacy" AND intervention)) OR ALL(("health literacy" AND interventions)) OR ALL("health literacy program") OR ALL("health literacy programs") OR ALL(("health literacy" AND program)) OR ALL(("health literacy" AND programs)) OR ALL("health literacy activity") OR ALL("health literacy activities") OR ALL(("health literacy" AND activity)) OR ALL(("health literacy" AND activities)) OR ALL("health literacy training") OR ALL("health literacy trainings") OR ALL(("health literacy" AND training)) OR ALL(("health literacy" AND trainings)) OR ALL((Education AND "oral health")) OR ALL("oral health education") OR ALL((workshop AND "oral health")) OR ALL("oral health workshop") OR ALL((workshops AND "oral health")) OR ALL("oral health workshops") OR ALL((workshop AND dental)) OR ALL((workshop AND "dental health")) OR ALL("Dental Health workshop") OR ALL((workshops AND "dental health")) OR ALL("Dental Health workshops") OR ALL((Education AND "dental health")) OR ALL("Dental Health Education") OR ALL("oral health program") OR ALL("dental health program") OR ALL((program AND "oral health")) OR ALL((program AND "dental health")) OR ALL("oral health training") OR ALL("dental health training") OR ALL((training AND "oral health")) OR ALL((training AND "dental health")) OR ALL("oral health activity") OR ALL("dental health activity") OR ALL("oral health activities") OR ALL("dental health activities") OR ALL((activity AND "oral health")) OR ALL((activity AND "dental health")) OR ALL((activities AND "oral health")) OR ALL((activities AND "dental health")) OR ALL("oral health intervention") OR ALL("oral health interventions") OR ALL("dental health intervention") OR ALL("dental health interventions") OR ALL("health program") OR ALL("health training") OR ALL("health activity") OR ALL("health programs") OR ALL("health workshop") OR ALL("health |
|  | workshops”) OR ALL(“health trainings”) OR ALL(“health activities”) OR ALL(“health intervention”) OR ALL(“health interventions”) OR ALL(“Patient Education as Topic”) OR ALL((Education AND Patient)) OR ALL(“Patient Education”) OR ALL(“Education of Patients”) AND (ALL(“oral health literacy”) OR ALL(“dental health literacy”) OR ALL(“literacy in dentistry”) OR ALL(“dental health literate”) OR ALL(“oral health literate”) OR ALL((“health literacy” AND “oral health”)) OR ALL((“health-related literacy” AND “oral health”)) OR ALL((“health Literacy” AND “dental health”))) |
| Google scholar | (“health education” OR (Education AND Health) OR “Community Health Education” OR Workshops OR Workshop OR “Training program” OR “Training programs” OR “training programme” OR (Training AND program) OR (Training AND programs) OR (training AND programme) OR “Educational activities” OR “Educational activity” OR (Educational AND activities) OR (Educational AND activity) OR “Educational workshop” OR (Educational AND workshop) OR “Educational workshops” OR (Educational AND workshops) OR “Educational program” OR “Educational programs” OR “Educational training” OR “Educational trainings” OR (Educational AND training) OR “Literacy program” OR “Literacy programs” OR (Literacy AND program) OR (Literacy AND programs) OR “health literacy workshop” OR “health literacy workshops” OR (“health literacy” AND workshop) OR (“health literacy” AND workshops) OR “health educational intervention” OR “health educational interventions” OR “health educational workshop” OR “health educational workshops” OR “health educational program” OR “health educational programs” OR “health educational activity” OR “health educational activities” OR “health educational training” OR “health educational trainings” OR “health education intervention” OR “health education interventions” OR “health education workshop” OR “health education workshops” OR “health education program” OR “health education programs” OR “health education activity” OR “health education activities” OR “health education training” OR “health education trainings” OR “health literacy intervention” OR “health literacy interventions” OR (“health literacy” AND intervention) OR (“health literacy” AND interventions) OR “health literacy program” OR “health literacy programs” OR (“health literacy” AND program) OR (“health literacy” AND programs) OR “health literacy activity” OR “health literacy activities” OR (“health literacy” AND activity) OR (“health literacy” AND activities) OR “health literacy training” OR “health literacy trainings” OR (“health literacy” AND training) OR (“health literacy” AND trainings) OR (Education AND “oral health”) OR “oral health education” OR (workshop AND “oral health”) OR “oral health workshop” OR (workshops AND “oral health”) OR “oral health workshops” OR (workshop AND dental) OR (workshop AND “dental health”) OR “Dental Health workshop” OR (workshops AND “dental health”) OR “Dental Health workshops” OR (Education AND “dental health”) OR “Dental Health Education” OR “oral health program” OR “dental health program” |
|  | OR (program AND “oral health”) OR (program AND “dental health”) OR “oral health training” OR “dental health training” OR (training AND “oral health”) OR (training AND “dental health”) OR “oral health activity” OR “dental health activity” OR “oral health activities” OR “dental health activities” OR (activity AND “oral health”) OR (activity AND “dental health”) OR (activities AND “oral health”) OR (activities AND “dental health”) OR “oral health intervention” OR “oral health interventions” OR “dental health intervention” OR “dental health interventions” OR “health program” OR “health training” OR “health activity” OR “health programs” OR “health workshop” OR “health workshops” OR “health trainings” OR “health activities” OR “health intervention” OR “health interventions” OR “Patient Education as Topic” OR (Education AND Patient) OR “Patient Education” OR “Education of Patients”) AND (“oral health literacy” OR “dental health literacy” OR “literacy in dentistry” OR “dental health literate” OR “oral health literate” OR (“health literacy” AND “oral health”) OR (“health-related literacy” AND “oral health”) OR (“health Literacy” AND “dental health”))<br>1228 |

## Appendix 2: Risk of bias assessment in randomized trial studies

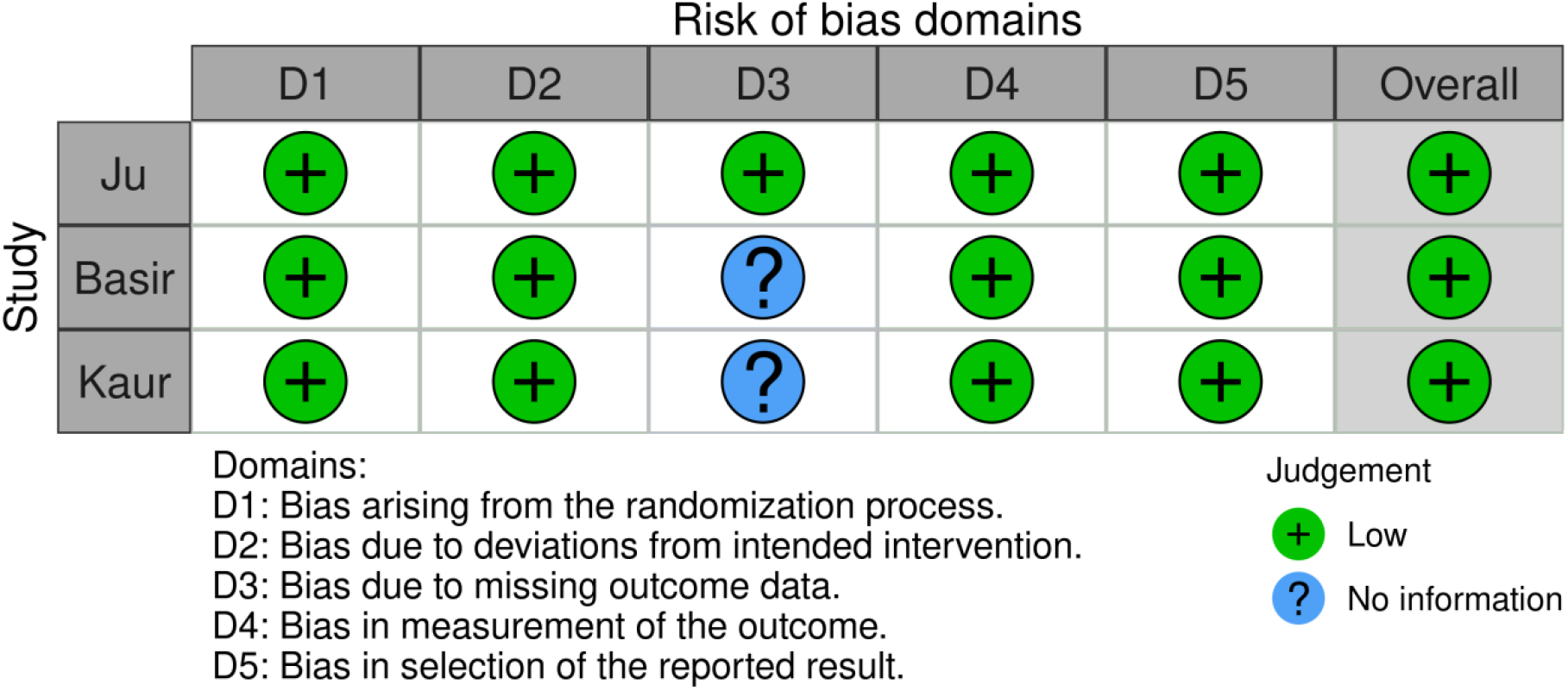

## Appendix 3. JBI Checklist for Quasi-Experimental Studies

| Checklist Question | Muller | Ismail | Farokhi, | Hjertstedt | sun |
| --- | --- | --- | --- | --- | --- |
| 1. Is it clear in the study what is the cause and what is the effect (i.e., there is no confusion about which variable comes first)? | Y | Y | Y | Y | Y |
| 2. Were the participants included in any comparisons similar? | Y | Y | Y | Y | Y |
| 3. Were the participants included in any comparisons receiving similar treatment/care, other than the exposure or intervention of interest? | Y | Y | Y | Y | Y |
| 4. Was there a control group? | N | Y | N | N | N |
| 5. Were there multiple measurements of the outcome both pre and post the intervention/exposure? | Y | Y | Y | Y | Y |
| 6. Was follow up complete and if not, were differences between groups in terms of their follow up adequately described and analyzed? | Y | Y | Y | Y | Y |
| 7. Were the outcomes of participants included in any comparisons measured in the same way? | Y | Y | Y | Y | Y |
| 8. Were outcomes measured in a reliable way? | Y | Y | Y | Y | Y |
| 9. Was appropriate statistical analysis used? | Y | Y | Y | Y | Y |
| Total | 8 | 9 | 8 | 8 | 8 |

Y: Yes N: No U: Unclear

